# Generalized logistic growth modeling of the COVID-19 outbreak in 29 provinces in China and in the rest of the world

**DOI:** 10.1101/2020.03.11.20034363

**Authors:** Ke Wu, Didier Darcet, Qian Wang, Didier Sornette

## Abstract

**Background:** the COVID-19 has been successfully contained in China but is spreading all over the world. We use phenomenological models to dissect the development of the epidemics in China and the impact of the drastic control measures both at the aggregate level and within each province. We use the experience from China to analyze the calibration results on Japan, South Korea, Iran, Italy and Europe, and make future scenario projections.

**Methods:** we calibrate the logistic growth model, the generalized logistic growth model, the generalized growth model and the generalized Richards model to the reported number of infected cases from Jan. 19 to March 10 for the whole of China, 29 provinces in China, four severely affected countries and Europe as a whole. The different models provide upper and lower bounds of our scenario predictions.

**Results:** We quantitatively document four phases of the outbreak in China with a detailed analysis on the heterogenous situations across provinces. Based on Chinese experience, we identify a high risk in Japan with estimated total confirmed cases as of March 25 being 1574 (95% CI: [880, 2372]), and 5669 (95% CI: [988, 11340]) by June. For South Korea, we expect the number of infected cases to approach the ceiling, 7928 (95% CI: [6341, 9754]), in 20 days. We estimate 0.15% (95% CI: [0.03%, 0.30%]) of Italian population to be infected in a positive scenario. We would expect 114867 people infected in Europe in 10 days, in a negative but probable scenario, corresponding to 0.015% European population.

**Conclusions:** The extreme containment measures implemented by China were very effective with some instructive variations across provinces. For other countries, it is almost inevitable to see the continuation of the outbreak in the coming months. Japan and Italy are in serious situations with no short-term end to the outbreak to be expected. There is a significant risk concerning the upcoming July 2020 Summer Olympics in Tokyo. Iran’s situation is highly uncertain with unclear and negative future scenarios, while South Korea is approaching the end of the outbreak. Both Europe and the USA are at early stages of the outbreak, posing significant health and economic risks to the world in absence of serious measures.

## 1. Background

Starting from Hubei province in China, the novel coronavirus (COVID-19) has been spreading all over the world, after two months of outbreak in China. Facing uncertainty and irresolution in December 2019 and the first half of January 2020, China then responded efficiently and massively to this new disease outbreak by implementing unprecedent containment measures to the whole country, including lockdown the whole province of Hubei and putting most of other provinces in de-facto quarantine mode. As of March 10, one and a half month after the national battle against the COVID-19 epidemic, China has successfully contained the virus transmission within the country, with new daily confirmed cases in mainland China excluding Hubei in the single digit range, and with just double digit numbers in Hubei. In contrast, many other countries have fast increasing numbers of confirmed cases. As of March 10, 103 countries in addition to China have reported confirmed cases infected by COVID-19.

A lot of efforts have been made in estimating the basic reproduction number R_0_ and predict the future trajectory of the coronavirus (COVID-2019) outbreak in the first quarter of 2020. In this paper, we focus on using phenomenological models without detailed microscopic foundations, but which have the advantage of allowing simple calibrations to the empirical reported data and providing transparent interpretations. This simple and top-down method can provide straightforward insights regarding the status of the epidemics and future scenarios of the outbreak. Usually, an epidemic follows an exponential growth at an early stage (following the law of proportional growth), peaks and then the growth rate decays as countermeasures to hinder the transmission of the virus are introduced.

There have been quite an extensive literature reporting statistical analysis and future scenarios based on phenomenological models. Most of previous work use simple exponential growth models and focus on the early growing process [1–4]. On the other hand, there are also many works arguing that the number of infected people follows a trajectory different from a simple exponential growth [5–15].

In this paper, we employ the logistic growth model, the generalized logistic growth model, the generalized growth model and the generalized Richards model, which have been successfully applied to describe previous epidemics [16–20]. All these models have some limitations and are only applicable in some stages of the outbreak, or when enough data points are available. Thus, we first calibrate different models to the reported number of infected cases in the COVID-19 epidemics from Jan. 19 to March 10 for the whole of China and 29 provinces in mainland China, and then draw some lessons useful to interpret the results of a similar modeling exercise performed on the four countries that are undergoing major outbreaks of this virus: Japan, South Korea, Iran, and Italy. Our analysis dissects the development of the epidemics in China and the impact of the drastic control measures both at the aggregate level and within each province. Borrowing from the experience of China, we made projections on the development of the outbreak in the four key countries and the whole Europe, based on different scenarios provided by the results from different models. Our study employs simple models to quantitatively document the effects of the Chinese containment measures against the COVID-19, and provide informative implications for the coming pandemic.

## 2. Data

### Confirmed cases

In this report, we focus on the daily data of confirmed cases in provinces in mainland China. We exclude the epicenter province, Hubei, which had a significant issue of underreporting at the early stage and also data inconsistency during mid-Feb due to a change of classification guidelines. For the provinces other than Hubei, the data is consistent except for one special event on Feb 20 concerning the data coming from several prisons. The data source is the national and provincial heath commission. For international data, the source is WHO. Note that the cases of the Diamond Princess cruise are excluded from Japan, following the WHO standard.

### Data adjustment

On Feb 20, for the first time, infected cases in the Chinese prison system were reported, including 271 cases from Hubei, 207 cases from Shandong, 34 cases from Zhejiang. These cases were concealed before because the prison system was not within the coverage of each provincial health commission system. Given that the prison system is relatively independent and the cases are limited, We remove these cases in our data for the modelling analysis to ensure consistency.

### Migration data

the population travels from Hubei and Wuhan to other provinces from Jan 1^st^ to Jan 23^rd^ are retrieved from the Baidu Migration Map (http://qianxi.baidu.com).

## 3. Method

We use the generalized Richards model (GRM), which is an extension of the original Richards growth model [21]. With three free parameters, the Richards growth model has been fitted to a range of logistic-type epidemic curves [16]. The generalized Richards model is defined by the differential equation:

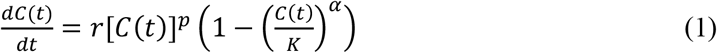

where *C(t)* represents the cumulative number of cases at time *t, r* is the growth rate at the early stage, and *K* is the final epidemic size. *p* ∈ [0,1] is a parameter that allows the model to capture different growth profiles including the constant incidence (*p* = 0), sub-exponential growth (0 < *p* < 1) and exponential growth (*p* = 1). The exponent α measures the deviation from the symmetric s-shaped dynamics of the simple logistic curve. The model recovers the original Richards model for *p* = 1, and reduces to the generalized logistic model [17] for α = 1 and *p* = 1.

For the calibration, we use the standard Levenberg–Marquardt algorithm to solve the non-linear least square optimization. For the GRM, the initial point C(0) ≔ C_0_ is fixed at the empirical value, and there are 4 remaining parameters (*K, r, p, α*) to be determined. For the fitting of the standard logistic growth function (*p* = 1 and α = 1), we free the initial point C_0_ and allow it being one of the 3 parameters to be optimized, as the early stage growth does not follow a logistic growth.

To estimate the uncertainty of our model estimates, we use a bootstrap approach with a negative binomial error structure NB(*μ*, σ^2^), where *μ* and σ^2^ are the mean and variance of the distribution, estimated from the empirical data.

## 4. Analysis at the global and provincial level for China (excluding Hubei)

### 4.1 Analysis at the aggregate level of mainland China (excluding Hubei)

As of March 10^th^, 2020, there are in total 13172 infected cases reported in the 30 provinces in mainland China outside Hubei. The initially impressive rising statistics has given place to a tapering associated with the limited capacity for transmission, exogenous control measure, and so on. In Figure 1, the trajectory of the total confirmed cases, the daily increase of confirmed cases, and the daily growth rate of confirmed cases in whole China excluding Hubei province are presented. The fits with the generalized Richards model and with the standard logistic growth model are shown in red and blue lines respectively in the upper, middle and lower left panel, with the data up to March 1^st^, 2020. In the lower left panel of Figure 1, the daily empirical growth rate 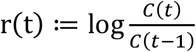 of the confirmed cases is plotted in log scale against time. We can observe two exponential decay regimes of the growth rate with two different decay parameters before and after Feb-14, 2020. The green line is the fitted linear regression line (of the logarithm of the growth rate as a function of time) for the data from Jan-25 to Feb 14, 2020, yielding an exponential decay parameter equal to −0.157 per day (95% CI: (−0.164, −0.150)). This indicates that, after the lockdown of Wuhan city on Jan 23 and the top-level health emergency activated in most provinces on Jan 25, the transmission in provinces outside Hubei has been contained with a relatively fast exponential decay of the growth rate from a value starting at more than 100% to around 2% on Feb 14. Then, starting Feb 15, three weeks after a series of extreme controlling measures, the growth rate is found to decay with a faster rate with a decay parameter equal to −0.277 per day (95% CI: (−0.313, −0.241)).

**Figure 1.**
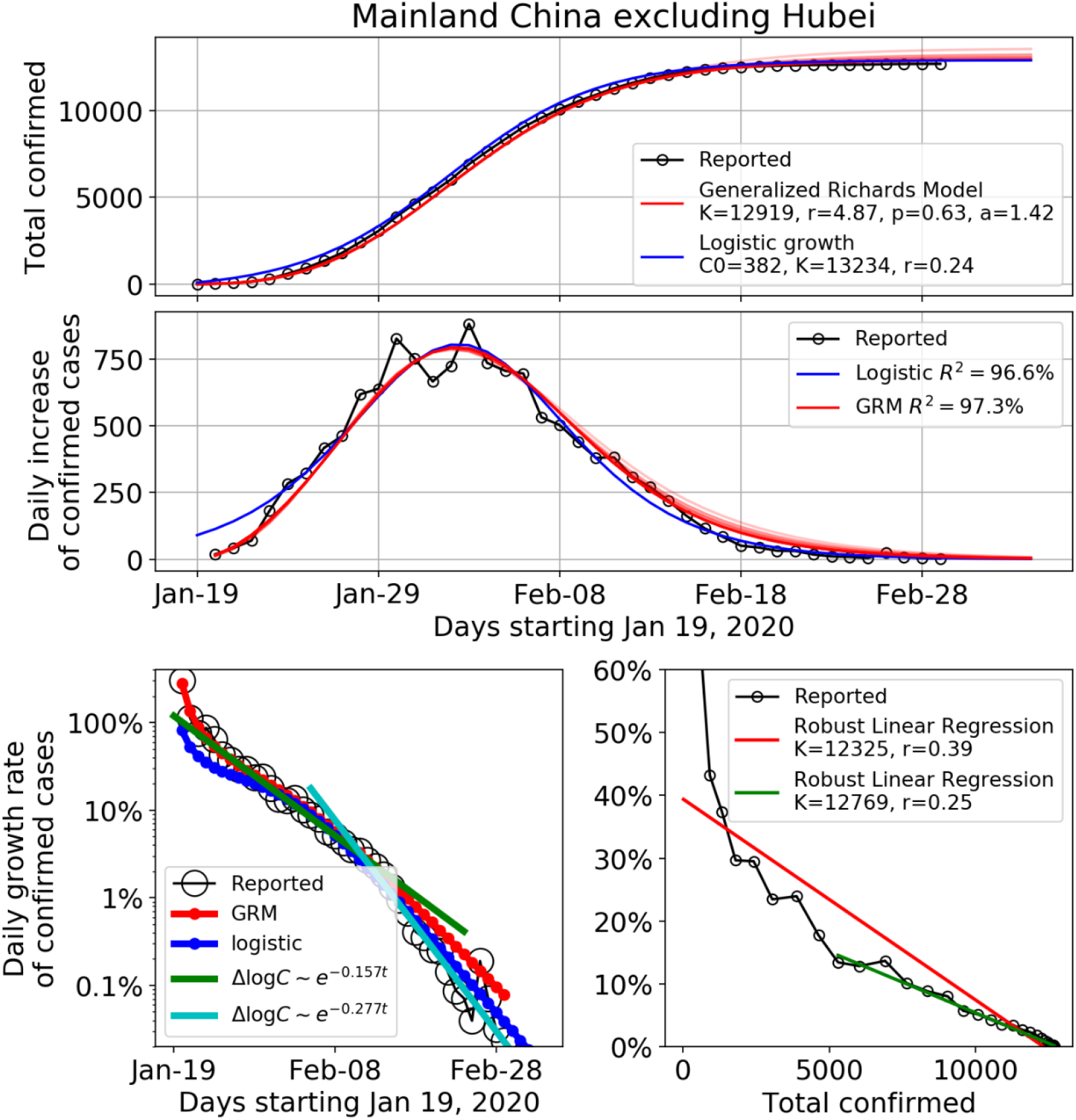
Time dependence of the total number of confirmed cases (upper panel), the daily number of new confirmed cases (middle panel), and the daily growth rate of confirmed cases (lower panel) in the mainland China excluding Hubei province until March 1^st^, 2020. The empirical data is marked by the empty circles. The blue and red lines in the upper, middle and lower left panels show the fits with the Logistic Growth Model and Generalized Richards Model (GRM) respectively. For GRM, we also show the fits using data ending 20, 15, 10, 5 days earlier than March 1^st^, 2020, as lighter red lines in the upper and middle panel. This demonstrates the consistency and robustness of the fits. The lower left panel shows the daily growth rate of the confirmed cases in log scale against time. The green and cyan straight lines show the linear regression of the logarithm of the growth rate as a function of time for the period of Jan 25 to Feb 14, and the period of Feb 15 to Mar 1, respectively. The lower right panel is the daily growth rate of the confirmed cases in linear scale against the cumulative number of confirmed cases. The red and green lines are the linear fits for the period of Jan 19 to Feb 1, and the period of Feb 2 to Mar 1, respectively.

This second regime is plotted as the cyan line in the lower left panel of Fig 1. The green and cyan straight lines show the linear regression of the logarithm of the growth rate as a function of time for the period of Jan 25 to Feb 14, and the period of Feb 15 to Mar 1, respectively. The asymptotic exponential decay of the growth rate can be justified theoretically from the generalized Richards model (1) by expanding it in the neighborhood where *C* converges to *K*. Introducing the change of variable *C*(*t*) = *K* (1 − *ε*(*t*)), and keeping all terms up to first order in *ε*(*t*), equation (1) yields

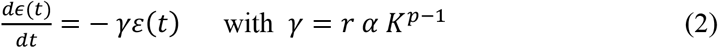

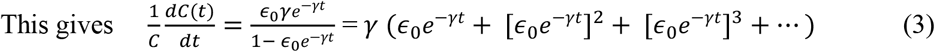

where *ε*_0_ is a constant of integration determined from matching this asymptotic solution with the non-asymptotic dynamics far from the asymptote. Thus, the leading behavior of the growth rate at long times is 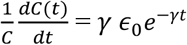, which is exponential decaying as shown in the lower left panel of figure 1. Using expression (2) for *γ* as a function of the 4 parameters *r, α, K* and p given in the inset of the top panel of figure 1, we get *γ* = 0.21 for mainland China excluding Hubei, which is bracketed by the two fitted values 0.17 and 0.28 of the exponential decay given in the inset of the lower left panel of figure 1.

In the lower right panel, the empirical growth rate *r*(*t*) is plotted in linear scale against the cumulative number of confirmed cases. The red and green lines are the linear regressed lines for the full period and for the period after Feb 1^st^, 2020 respectively. We can see that the standard logistic growth cannot capture the full trajectory until Feb 1^st^. After Feb 1^st^, the linear fit is good, qualifying the simple logistic equation (*p* = 1 and α = 1), with growth rate estimated as r=0.25 for the slope, which is compatible with the value determined from the calibration over the full data set shown in the top two panels of figure 1.

Figure 2 demonstrates the sensitivity of the calibration of the GRM to the end date of the data by presenting six sets of results for six end dates. Specifically, the data on the daily number of new confirmed case is assumed to be available until 23 Jan, 28 Jan, 2 Feb, 7 Feb, 12 Feb, 17 Feb, i.e. 30, 25, 20, 15, 10 and 5 days before Feb 22 were presented. For each of the six data sets, we generated 500 simulations of 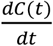 based on the best fit parameters using parametric bootstrap with a negative binomial error structure, as in prior studies [17]. Each of these 500 simulations constitutes a plausible scenario for the daily number of new confirmed cases, which is compatible with the data and GRM. The dispersion among these 500 scenarios provides a measure of stability of the fits and their range of values gives an estimation of the confidence intervals. The first conclusion is the non-surprising large range of scenarios obtained when using data before the maximum, which however encompass the realized data. We observe a tendency for early scenarios to predict a much faster and larger number of new cases than observed, which could be expected in the absence of strong containment control. With more data, the scenarios become more accurate, especially when using realized data after the peak, and probably account now well for the impact of the containment measures that modified the dynamics of the epidemic spreading.

**Figure 2.**
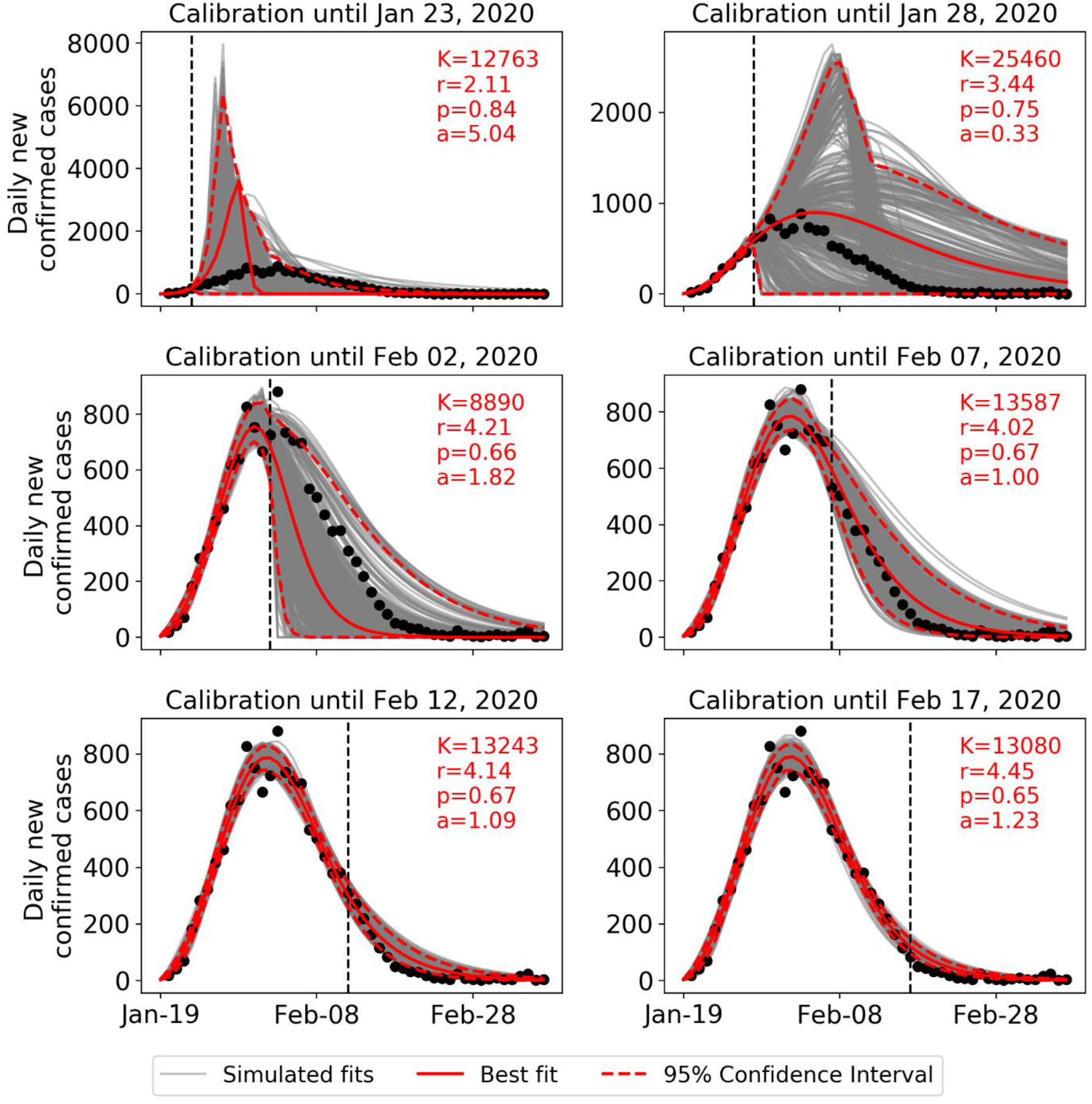
Daily number of new observed confirmed cases for mainland China excluding Hubei (black circles) compared with 500 scenarios built by parametric bootstrap with a negative binomial error structure on the GRM model with best fit parameters determined on the data up to the time indicated by the vertical dashed line. The last time used in the calibration is respectively 5, 10, 15, 20, 25, 30 days before Feb 22, 2020 from bottom to top. The red continuous line is the best fitted line and the two dashed red curves delineate the 95% confidence interval extracted from the 500 scenarios. The six panels correspond each to a different end date, shown as the sub-title of each panel, at which the data has been calibrated with the GRM model.

### 4.2 Analysis at the provincial level (29 provinces) of mainland China (excluding Hubei)

As of March 1, 2020, the daily increase of the number of confirmed cases in China excluding Hubei province has decreased to less than 10 cases per day. The preceding one-month extreme quarantine measures thus seems to have been very effective from an aggregate perspective. At this time, it is worthwhile to take a closer look at the provincial level to study the effectiveness of measures in each province. The supplementary material presents figures similar to figure 1 for each of the 29 provinces in mainland China. Tibet is excluded as it only has 1 confirmed case as of March 1. Table 1 provides some useful statistics for each province and the values of the fitted parameters of the generalized Richards model, logistic growth model and the exponential decay exponent of the growth rate. This analysis at the 29 provinces allows us to identify four phases in the development of the epidemic outbreak in mainland China.

**Table 1:**
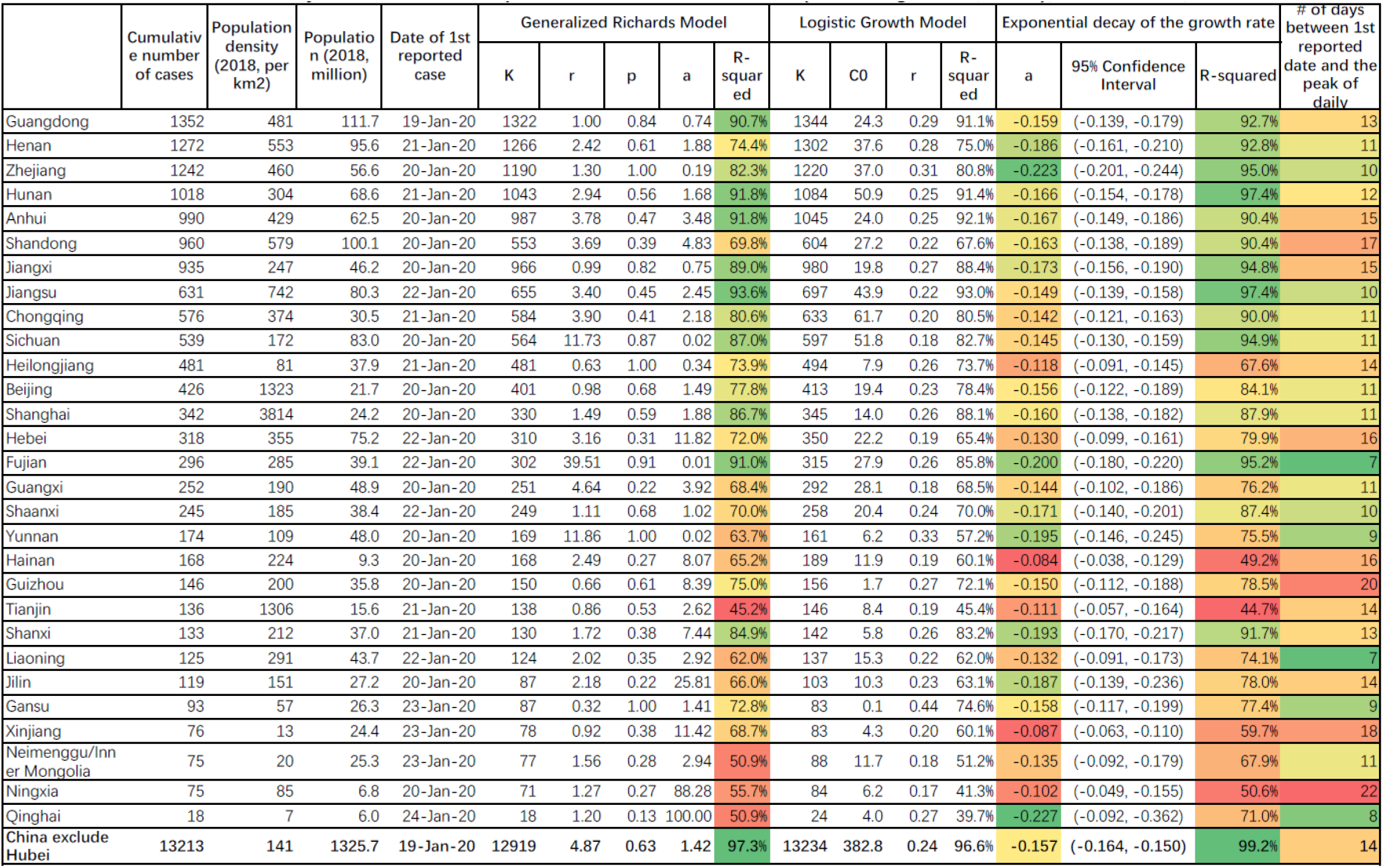
Summary statistics for 29 provinces in mainland China (excluding Hubei and Tibet), as of Feb 29, 2020. The values of the fitted parameters of the generalized Richards model, logistic growth model and the exponential decay exponent are reported.

#### Phase I (Jan 19 – Jan 24, 6 days): early stage outbreak

The data mainly reflects the situation before Jan 20, when no measures were implemented, or they were of limited scope. On Jan 19, Guangdong became the first province to declare a confirmed case outside Hubei in mainland China [22]. On Jan 20, with the speech of President Xi, all provinces started to react. As of Jan 24, 28 provinces reported confirmed cases with daily growth rates of confirmed cases ranging from 50% to more than 100%.

#### Phase II (Jan 25 – Feb 1, 8 days): fast growth phase approaching the peak of the incidence curve (inflection point of the cumulative number)

The data starts to reflect the measures implemented in the later days of Phase I and in Phase II. In this phase, the government measures against the outbreak have been escalated, marked by the lockdown of Wuhan on Jan 23, the top-level public health emergency state declared by 20+ provinces by Jan 25, and the standing committee meeting on Jan 25, the first day of the Chinese New Year, organized by President Xi, to deploy the forces for the battle against the virus outbreak. In this phase, the growth rate of the number of confirmed cases in all provinces are declining from 50% to 10%+, with an exponentially decay rate of 0.157 for the aggregated data. At the provincial level, some provinces failed to see a continuous decrease of the growth rate and witnessed the incidence grow at a constant rate for a few days, implying exponential growth of the confirmed cases. These provinces include Jiangxi (∼40% until Jan 30), Heilongjiang (∼25% until Feb 5), Beijing (∼15% until Feb 3), Shanghai (∼20% until Jan 30), Yunnan (∼75% until Jan 27), Hainan (∼10% until Feb 5), Guizhou (∼25% until Feb 1), Jilin (∼30% until Feb 3). Some other provinces managed to decrease the growth rate exponentially during this period. As of Feb 1^st^, 15 provinces had reached the peak of the incidence curve, indicating the effectiveness of the extreme measures, and most provinces started to be in control of the epidemics.

#### Phase III (Feb 2 – Feb 14, 13 days): slow growth phase approaching the end of the outbreak

In this period, all provinces continued to implement their strict measures, striving to bring the epidemics to an end. The growth rate of the number of confirmed cases declined exponentially with similar rates as in Phase II, pushing down the growth rate from 10% to 1%. In phase III, all provinces have passed the peak of the incidence curve, which allows us to obtain precise scenarios for the dynamics of the end of the outbreak from the model fits (Figure 2). As of Feb 14, 23 out of 30 provinces have less than 10 new cases per day.

#### Phase IV (Feb 15 – 8 March): the end of the outbreak

Starting Feb 15, the exponential decay of the growth rate at the aggregate level has switched to an even faster decay with parameter of 0.277 (Figure 1). As of Feb 17, one week after normal work being allowed to resume in most provinces, 22 provinces have a growth rate smaller than 1%. As of Feb 21, 28 provinces have achieved 5-day average growth rates smaller than 1%.

## 5. Analysis of the development of the epidemic and heterogenous provincial responses

### 5.1 Quantification of the initial reactions and ramping up of control measures

On Jan 19, Guangdong was the first province to report a confirmed infected patient outside Hubei. On Jan 20, 14 provinces reported their own first case. During Jan 21-23, another 14 provinces reported their first cases. If we determine the peak of the outbreak from the 5 days moving average of the incidence curve, then there are 15 provinces taking 7-11 days from their first case to their peak, 9 provinces taking 12-15 days, and 6 provinces taking more than 15 days. If we define the end of the outbreak as the day when the 5 days moving average of the growth rate becomes smaller than 1%, then 7 provinces spent 8-12 days from the peak to the end, 7 provinces spent 13-16 days, 13 provinces spent 17-20 days, and 2 provinces spent 21-22 days. For the six provinces that have the longest duration from the start of their outbreak to the peak (more than 15 days), it took 8-13 days for them to see the end of the outbreak (Figure 3). This means that these 6 provinces were able to control the local transmissions of the imported cases quite well, so that the secondary transmissions were limited. In contrast, 20 provinces took 28-31 days from the start to the end of the outbreak. Thus, those provinces that seem to have responded sluggishly during the early phase of the epidemics seem to have ramped up aggressively their countermeasures to achieve good results.

**Figure 3.**
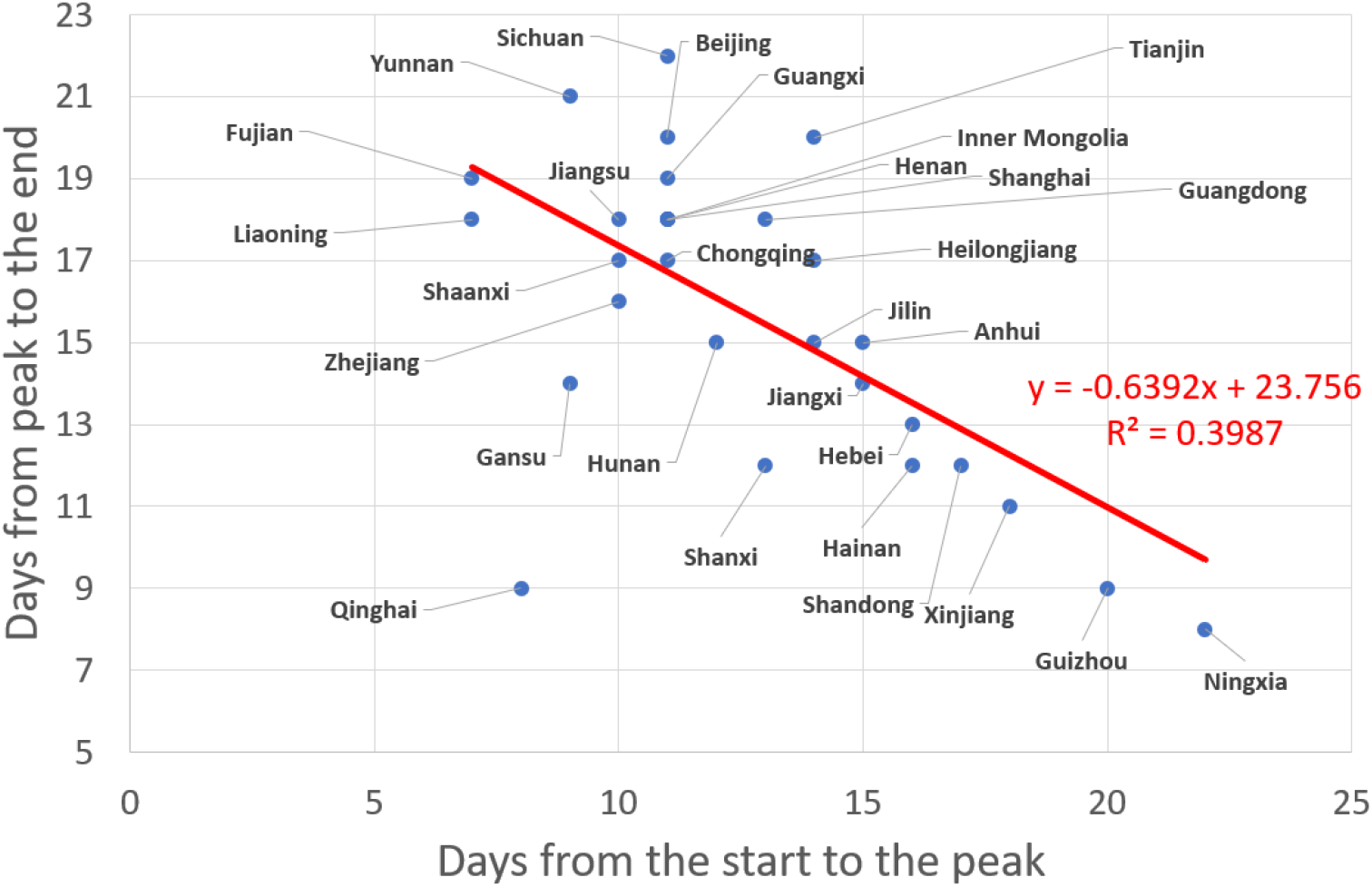
Inverse relationship found across the 29 Chinse provinces between the number of days from peak to the end and the duration from start to the peak of the epidemics. Here, the end of the outbreak is defined operationally as the day when the 5 days moving average of the growth rate becomes smaller than 1%.

### 5.2 Quality of fits with the logistic equation as proxy to early under-reporting

Both the generalized Richards model and the logistic growth model can capture most of the dynamics at the cumulative level, daily increase level (1^st^ derivative) and the daily growth rate level (2^nd^ derivative). However, likely due to the potential underreporting at the beginning, the logistic growth curve fails to capture the growth dynamics at the early stage in most provinces. The logistic growth provides a better quality of fit after Feb 1^st^ in general, as we can see from the lower right panel in the figure for each province (see supplementary material).

### 5.3 Diagnostic of the efficiency of control measures from the exponential decay of the growth rate of infected cases

The 10 most infected provinces (Guangdong, Henan, Zhejiang, Hunan, Anhui, Jiangxi, Shandong, Jiangsu, Chongqing, Sichuan) have done quite well in controlling the transmission, as indicated by the fact that their daily growth rates follow well-defined exponential decays, with a *R*^2^ larger than 90%. This exponential decay continued for all ten cities until the situation was completely under control during Feb 15-18, when the daily incidence was at near zero or a single-digit number. Eight out of these ten provinces have a decreasing parameter of the exponential decay of the growth rate ranging from 0.142 to 0.173, similar to what is observed at the national average level (0.157). Note that this exponential decay can be inferred from the generalized Richards model, as we noted in section 4.1.

### 5.4 Zhejiang and Henan exemplary developments

Zhejiang and Henan are the 2^nd^ and 3^rd^ most infected provinces but have the fastest decaying speed of the incidence growth rate (decay parameter for Zhejiang: 0.223, Hunan: 0.186) among the most infected provinces. This is consistent with the fast and strong control measures enforced by both provincial governments, which have been praised a lot on Chinese social network [23,24]. As one of the most active economies in China and one of the top provinces receiving travelers from Wuhan around the Lunar New Year [25], Zhejiang was the first province launching the top-level public health emergency on Jan 23^rd^, and implemented strong immediate measures, such as closing off all villages in some cities. The fitted curves from the GRM and logistic growth models indicate a peak of the incidence curve on Jan 31, which is the earliest time among top infected provinces. Similarly, Henan Province, as the neighbor province of Hubei and one of the most populated provinces in China, announced the suspension of passenger bus to and from Wuhan at the end of Dec 2019. In early Jan 2020, Henan implemented a series of actions including suspending poultry trading, setting up return spots at the village entrances for people from Hubei, listing designated hospitals for COVID-19 starting as early as Jan 17, and so on. These actions were the first to be implemented among all provinces.

### 5.5 Heterogeneity of the development of the epidemic and responses across various provinces

Less infected provinces exhibit a larger variance in the decaying process of the growth rate. However, we also see good examples like Shanghai, Fujian and Shanxi, which were able to reduce the growth rate consistently with a low variance. These provinces benefited from experience obtained in the fight against the 2003 SARS outbreak or enjoy richer local medical resources [7]. This enabled the government to identify as many infected/suspected cases as possible in order to contain continuously the local transmissions. Bad examples include Heilongjiang, Jilin, Tianjin, Gansu, which is consistent with the analysis of [7].

Most provinces have a small parameter of *p* of the GRM (see equation (1)) and an exponent α large than 1, indicating that China was successful in containing the outbreak as sub-exponential growing process (*p* < 1), with a faster than logistic decay (*α* > 1). Guangdong, Zhejiang, Jiangxi, Sichuan, Heilongjiang, Fujian, Yunnan and Gansu do not fall into this category of small *p* and large α. However, this does not necessarily indicate ineffective measures for these provinces. The large *p* and small α in Guangdong and Zhejiang are likely to be mainly due to high population densities and highly mobile populations in mega-cities, which are factors known to largely contribute to the fast transmission of viruses. Jiangxi, Sichuan, Fujian, Yunnan and Gansu all had a fast growth phase before Feb 1^st^, but were successful in controlling the subsequent development of the epidemics. The fast growth phase in Heilongjiang lasted a bit longer than the abovementioned provinces, due to the occurrence of more numerous local transmissions. Heilongjiang has been criticized a lot for its high infected cases and death rate (2.7%), given that it is far from Hubei and does not have a large number of migrating people from Hubei.

### 5.6 Initial and total confirmed numbers of infected cases correlated with travel index

The initial value C_0_ of the logistic equation could be used as an indicator of the early number of cases, reflecting the level of early contamination from Hubei province as the epicenter of the outbreak. To support this proposition, the upper panel of Figure 4 plots the estimated C_0_ as a function of the migration index from Hubei & Wuhan to each province. The migration index is calculated as equal to 25% of the population migrating from Hubei (excluding Wuhan) plus 75% of the population migrating from Wuhan, given that Wuhan was the epicenter and the risks from the Hubei region excluding Wuhan are lower. One can observe a clear positive correlation between the estimated C_0_ and the migration index. The lower panel of figure 4 shows an even stronger correlation between the total number of cases recorded on March 6^st^ and the travel index, expressing that a strong start of the epidemics predicts a larger number of cases, which is augmented by infections resulting from migrations out of the epidemic epicenter.

**Figure 4.**
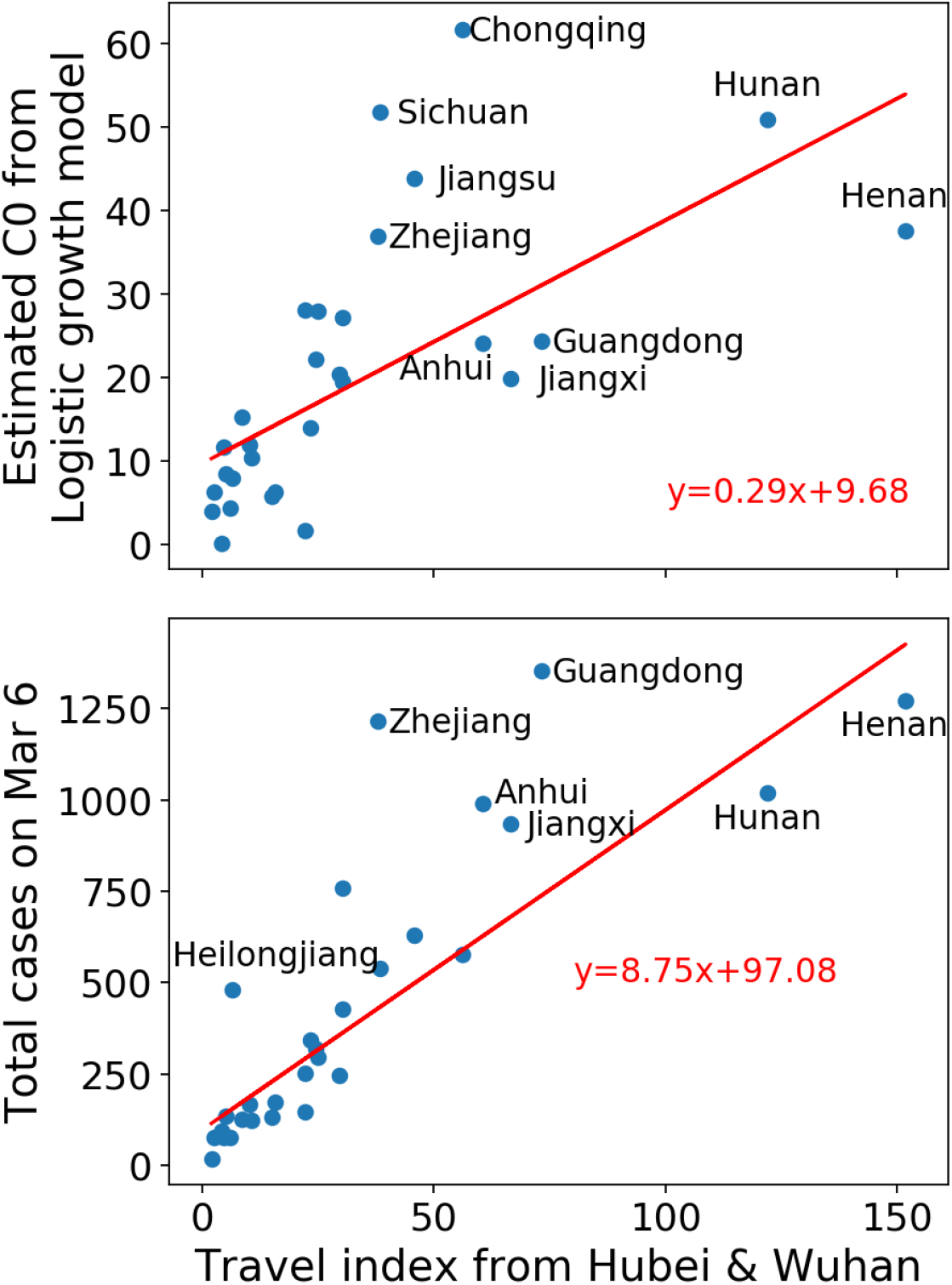
Upper panel: estimated C_0_ for the logistic growth model versus travel index from Hubei & Wuhan. Lower panel: total confirmed cases versus travel index from Hubei & Wuhan. The Pearson correlation between C_0_ and the migration index is 0.65 (p < 10^−3^), and the correlation between the cumulative number of confirmed cases and the migration index is 0.82 (p < 10^−4^).

## 6. Analysis of the recent development in Japan, South Korea, Iran and Italy and the world

Although it is clear that the virus in China has been successfully controlled up to the time of finalizing this paper, it has spread to 117 territories in the world as of March 10, with Italy, South Korea, Japan, and Iran as the four major hotspots. The highly connected European continent is at the beginning phase of the outbreak. In this section, we will analyze the situation in the four countries that have recently reported increasing numbers, and provide forecasts based on several models.

### 6.1 Models

Most of the countries analyzed here are still at an early stage of the outbreak, i.e., the cumulative number of cases is not yet reaching the inflection point. In this regime, the generalized Richards model, which has an additional parameter α describing the asymmetry between growth and decay of the incidence curve, is not sufficiently constrained by the limited available data. Therefore, we consider the following three simpler models with fewer parameters:

− Logistic growing model:

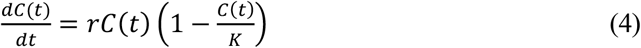

− Generalized Logistic model (GLM):

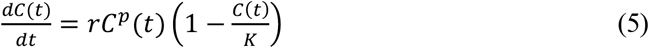

− Generalized growth model (GGM):

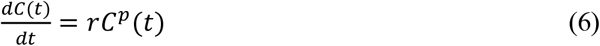

The generalized growth model (6) allows for a sub-exponential growth of the outbreak in the early stage (for p < 1), but cannot describe the decay of the incidence rate. It thus serves as a rough upper limit, obtained by assuming that the outbreak continues to grow following the same process as in the past. The generalized Logistic model (5) and Logistic growth model (4) both assume a logistic decay of the growth rate as the total number of confirmed cases increases. Note that they are nested in the generalized Richards model (1), by fixing respectively *α* = 1 and *α* = 1; p = 1. Both Logistic type models tend to under-estimate the total size of the infected population at the early stage, so they should provide lower bounds. The generalized Logistic model allows for an early sub-exponential growth and can better describe the possible asymmetry between growth and decay dynamics. We now calibrate these three models to the data of each of the four countries: Italy, South Korea, Japan, and Iran. The fitting procedure and simulation of confidence intervals are the same as for the generalized Richards model presented for the Chinese data sets in previous sections.

### 6.2 Results of the calibration of three growth models to the epidemic dynamics in Japan, South Korea, Iran. Italy and the world

#### 6.2.1 Japan

Among the four countries, Japan and South Korea are the two first countries that reported confirmed cases. However, the situations are very different, as reflected from the statistics of these two Asian countries. Japan has reacted quite fast after the confirmation of the human-to-human transmission of the COVID-19. On 24 January, Prime Minister Abe convened the “Ministerial Meeting on Countermeasures Related to the Novel Coronavirus” at the Prime Minister’s Office with members of his Cabinet. On 30 January, PM Abe announced the establishment of “The Novel Coronavirus Response Headquarters” and has been actively developing countermeasures against the virus outbreak. As of March 10, Japan has been reporting new cases consecutively since Feb 11, with a total of 567 confirmed cases, and a relatively stable daily growth rate at 11.8% (standard deviation 6.3%). Thanks to the early response of the Japanese government, there is no large-scale community transmission in Japan, and this growth rate is not high compared to the initial outbreak stages in China and many other countries.

However, as showed in Figure 5, we do not observe any decay of this growth rate, in contrast to what occurred in phase III in mainland China beyond the inflection point of the cumulative number of cases with a continuous exponential decrease of the growth rate from 10% to 1%. Figure. 5 shows that the number of confirmed cases is increasing exponentially. If this goes on, the total cumulative number of cases will reach 806 (95% CI: [683,957]) as of March 15, 1185 (95% CI: [932, 1603]) as of March 20, and 1701 (95% CI: [1219, 2716]) as of March 25, predicted by the generalized growth model.

**Figure 5.**
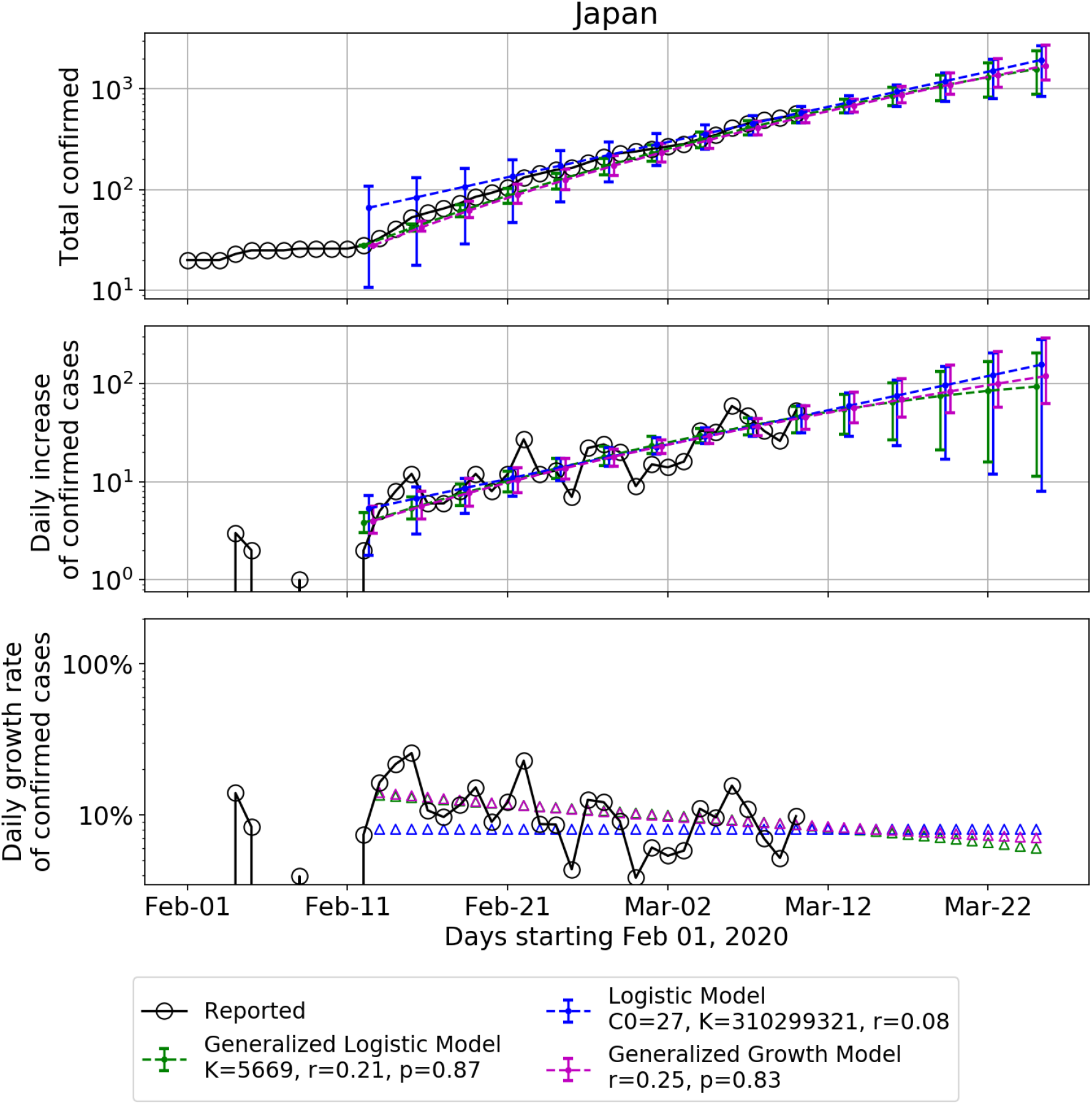
Calibration of three models (Generalized Logistic Model in green, Generalized Growth Model in purple and Logistic Model in blue) to data shown with empty circles for Japan. Upper panel: fitted and predicted cumulative confirmed cases. Middle panel: fitted and predicted daily incidence. The lower panel plots the daily growth rate. The fitted data is plotted every three days, and the error bar is the 95% confidence interval extracted from the 500 simulations assuming a negative binomial error structure. The fitting is based on data since Feb 12, 2020.

The parameter *p* of the GLM is estimated as 0.92 (95% CI: [0.73, 1]), and as 0.89 (95% CI: [0.63, 1.00]) for the GGM. This confirms that the growth is closed to exponential, indicating that local transmission channels in Japan are not yet under control. The quasi-exponential growth rationalizes the fact that parameter *K* for the logistic model is found to be so large, such that the quadratic term in equation (4) is essentially irrelevant.

In a positive scenario proxied by the generalized logistic model, allowing for sub-exponential growth and arriving at the inflection point in 24 days, the estimated total number of cases as of March 25 will be 1574 (95% CI: [880, 2372]), and 5669 (95% CI: [988, 11340]) by the end of May. Most cases in Japan are imported or cases with contact history to those imported, with no evidence of large-scale community transmission. This is similar to phase II-III in mainland China.

However, it took China two weeks to reduce the growth rate from 10% to 1% under extreme containment measures. Given that we do not expect the same level of measures can be implemented in Japan sooner or later, the future scenario for Japan is highly uncertain, and will depend on whether the government decides to increase the containment measures. Without stronger and fast measures introduced in Japan, we see a significant risk concerning the upcoming July 2020 Summer Olympics in Tokyo.

#### 6.2.2 South Korea

In contrast to Japan, the analysis of South Korea’s data indicates that the transmission is under control, although the total number of confirmed cases is much larger than Japan. This is likely due to the much larger efforts to test the population, with a ratio of close to 4 tests per 1000 inhabitants, compared with 0.066 tests per 1000 inhabitants for Japan.

Figure 6 shows that the dynamics of the infected population is clearly not an exponential growth, as the growth rate has decreased significant from 40% at the end of Feb to 1.8% on March 10. The first phase of the outbreak in South Korea occurs from Jan 20 to Feb 17, when most cases were imported or have a clear contact history to the imported cases. On 18 February, South Korea confirmed its 31st case in Daegu, a member of the Shincheonji religious organisation. The patient continued to attend gatherings of Shincheonji days after showing symptoms of the illness. Such gatherings are typically held with people in very close proximity and include physical contact between the members. This likely triggered the second phase of the outbreak and brought the total number of confirmed cases to 7513 as of March 10. Our models fit to the data starting from Feb 18.

**Figure 6.**
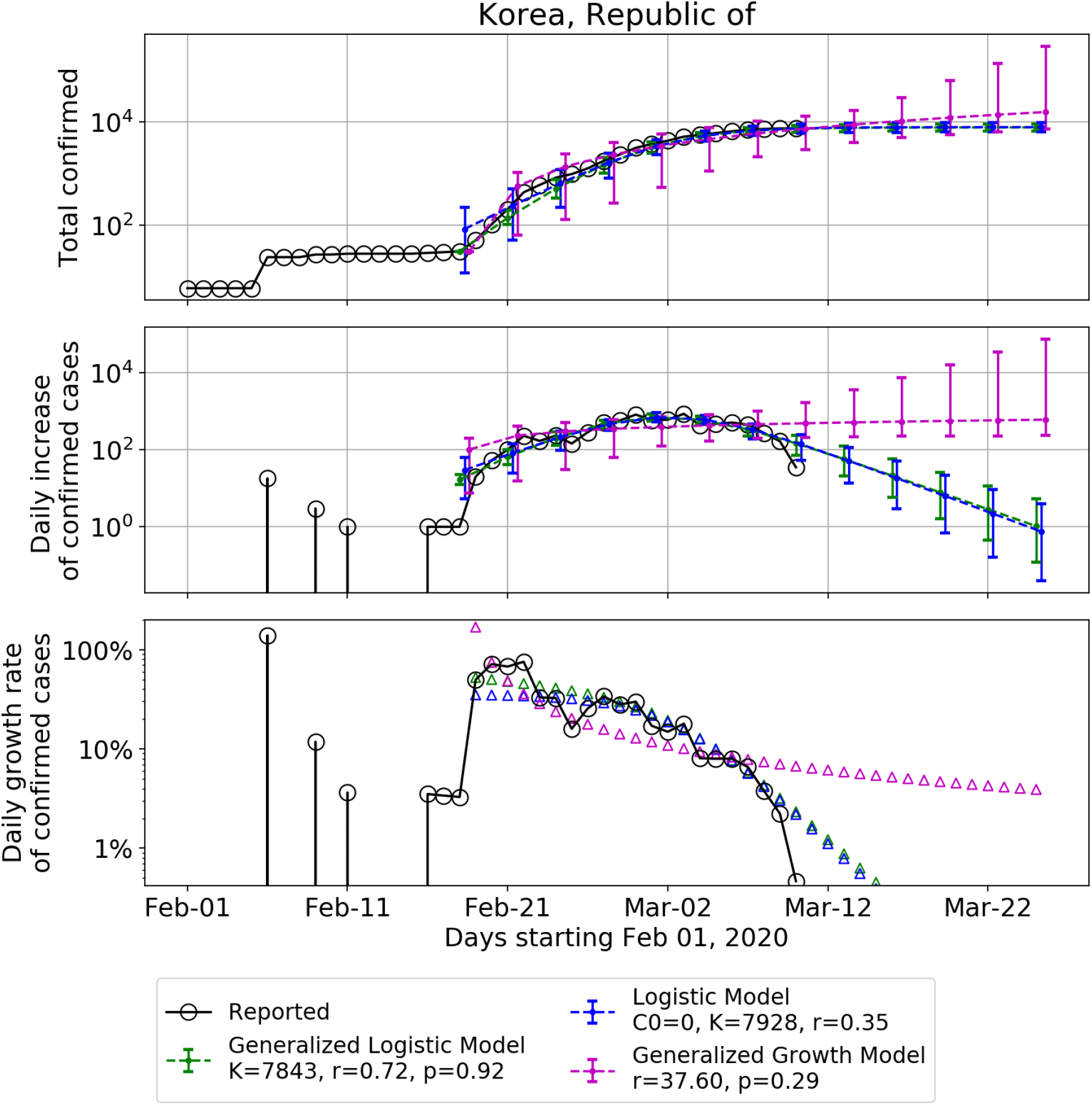
Same as figure 5 for South Korea. The fitting is based on data since Feb 18, 2020.

Figure 6 shows that the infection point has been passed in recent days, and the daily number of new cases is starting to go down. This is a good sign and we can expect the outbreak to follow the logistic type decay in subsequent days, assuming the absence of another large-scale community transmission, as in phase III of many provinces in China. Both the generalized Logistic model and Logistic model provide similar estimates of the final number of infected cases: 7843 (95% CI: [6594, 9138]) from GLM and 7928 (95% CI: [6341, 9754]) for the logistic model, both of which could be expected to be reached within 20 days. However, whether the daily incidence can decay symmetrically to the increase phase will depend largely on the strategy of the government. In China, it was mainly the extreme measures of quarantines that helped most provinces’ incidences to decay fast. The South Korean government has been actively tracing all possible cases and performing more than 10,000 tests per day as of May 8 [26], trying to cut off all the potential transmission sources.

#### 6.2.3 Iran

Compared to Japan and South Korea, Iran and Italy are currently in a worse situation. Iran reported its first confirmed cases on Feb 19. As of 10 March 2020, according to Iranian health authorities, there had been 291 deaths in Iran with a total of 8042 confirmed infections, indicating a death rate of 3.6%, conditional of being infected.

As showed in Figure 7, although the growth rate of the confirmed cases is decreasing, the future scenarios are still highly uncertain as the total number of infected cases is just around the inflection point. Plus, there may be new rounds of large-scale outbreaks. A possible scenario is that the growth rate may continue to float and fluctuate around the current rate for a while, for which the generalized growth model becomes relevant and can be used for extrapolation. In this scenario, the total number of cases would grow to 14994 (95% CI: [10432, 21048]) in 5 days (Mar-15), and to 23025 (95% CI: [15246, 42249]) in 10 days (Mar-20).

**Figure 7.**
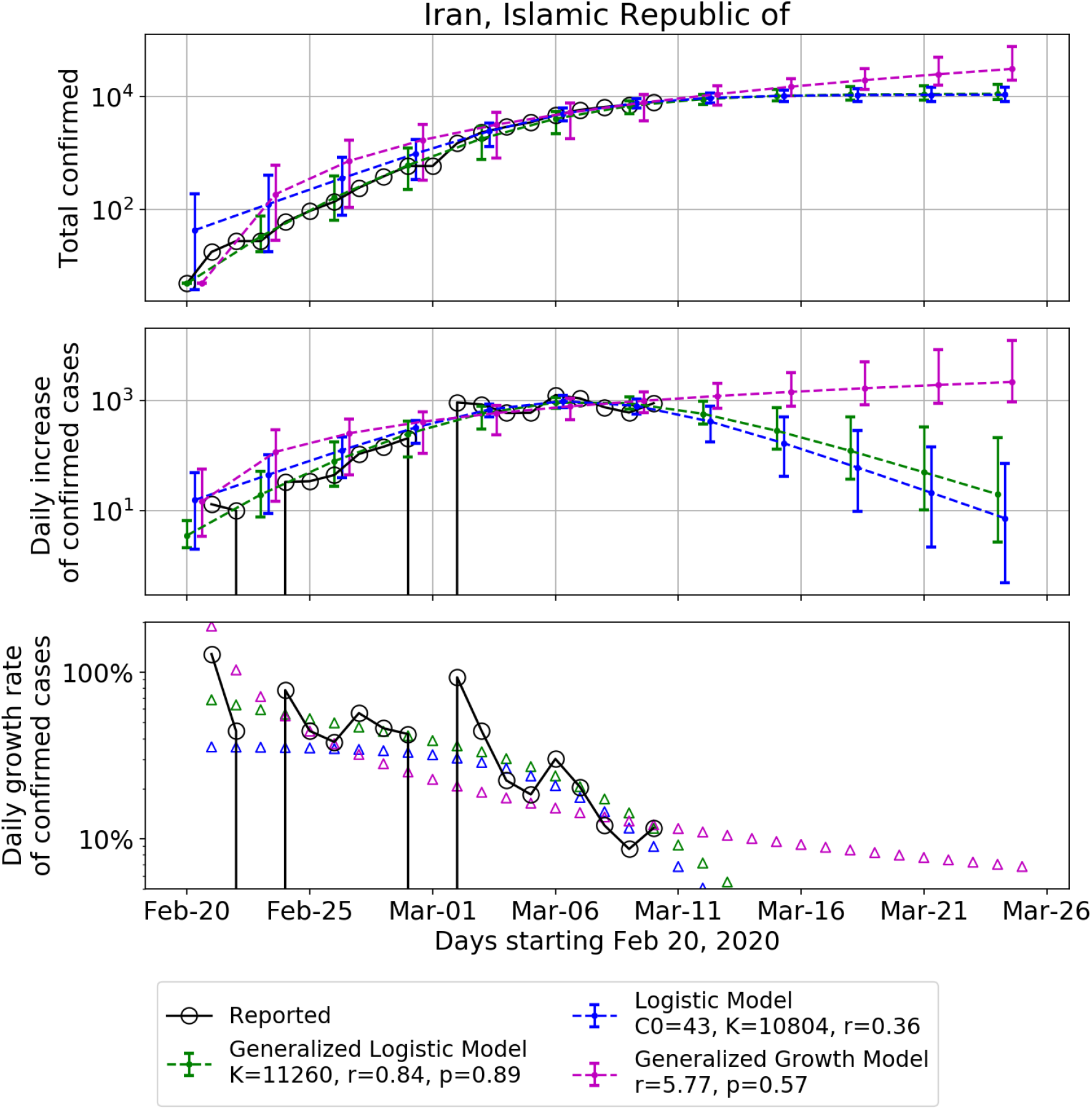
Same as figure 5 for Iran. The fitting is based on data since Feb 20, 2020.

On the other hand, a more positive scenario could be the one proposed by the logistic model, which provides an estimate of the total number of infected cases of 10719 (95% CI: [8178, 14390]) in 10 days (Mar-20), and a final number of 10804 cases (95% CI: [8202, 15060]) in 20 days. The results obtained from the generalized logistic model are similar to those of the logistic model, with a smaller variance. The belated response, lack of resources, and opaque information communication in Iran lead to large uncertainties regarding the future development of the epidemic in this country. It would be already a very positive outcome if the data could follow the two logistic models.

#### 6.2.4 Italy

The situation in Italy is quite severe at the moment of writing. As of March 10, Italy has the largest number of confirmed cases (9172) and deaths (463) among all the countries except China. The outbreak in Italy was mainly due to a few outbreak clusters, such as Lombardy and Veneto. Similar to Japan, the growth rate of the confirmed cases at Italy has been fluctuating between 20% and 30% for a week, which is still far from the inflection point.

The generalized logistic model provides the best scenario, i.e. predicting that the inflection point will be reached in 10 days. This scenario will already lead to a total cumulative number of infected cases of 59199 (95% CI: [16796, 107097]) in 15 days, and to 91719 (95% CI: [16944, 183440]) until the end of the outbreak.

In contrast, a pessimistic scenario provided by the Logistic model predicts that there could be up to 2 million infected cases adding up until the end of May.

In any case, the situation in Italy is predicted to become worse than at the epicenter province Hubei in China. Moreover, as Italy is a highly connected node in the global network, it poses significant risks to other countries. As of March 2, 30 countries and territories have confirmed cases which appear to have originated from Italy [27]. On Mar 8, the government of Italy imposed a quarantine on the region of Lombardy and fourteen neighboring provinces in Piedmont, Veneto, Emilia-Romagna, and Marche, in response to the growing outbreak of COVID-19 in the country, putting more than a quarter of the national population (ca. 16 million people) under lockdown. On March 10, Italy has extended its emergency coronavirus measures, which include travel restrictions and a ban on public gatherings, to the entire country. This will certainly help to reduce the transmission of the virus. However, given the experience in China, the effect will only be reflected in the statistics after a delay of at least one week, and it would be already a positive outcome if the outbreak would follow the path described the generalized logistic model.

#### 6.2.5 Europe and the world

Figure 9 shows the number of confirmed cases per million people in the four hotspot countries, in comparison with mainland China excluding Hubei, the epicenter Hubei province, Europe, Singapore, Hong Kong, and the USA. Although Hubei has the highest infection ratio above 0.1%, South Korea and Italy have become the most infected country in the world. The high statistics in South Korea is likely as mentioned above due to the largest number of performed tests. We should indeed stress that the actual cumulative number of infected cases is probably misleading as it is likely that the incidence is an increasing function of testing.

**Figure 8.**
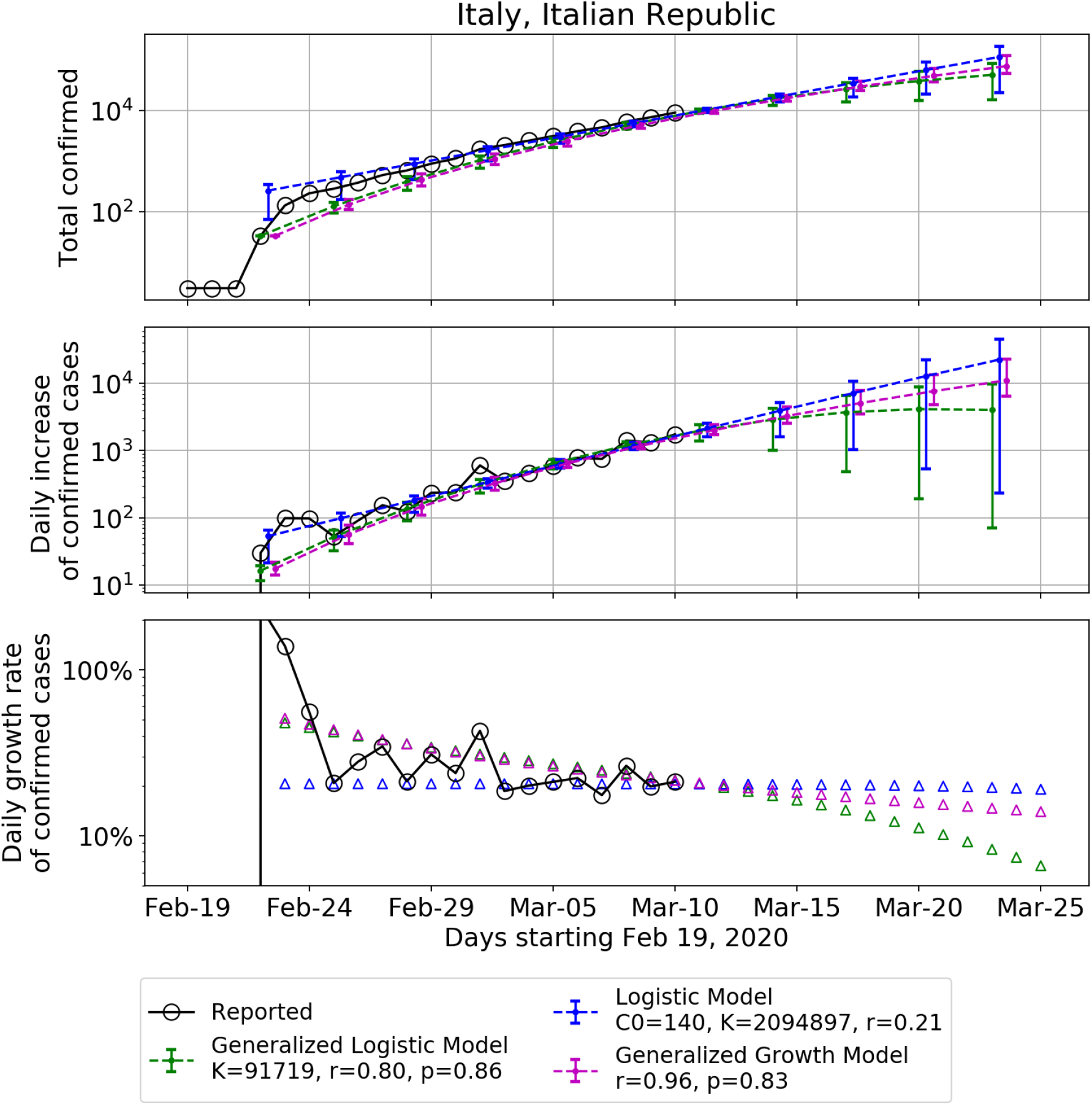
Same as figure 5 for Italy. The fitting is based on data since Feb 22, 2020.

**Figure 9.**
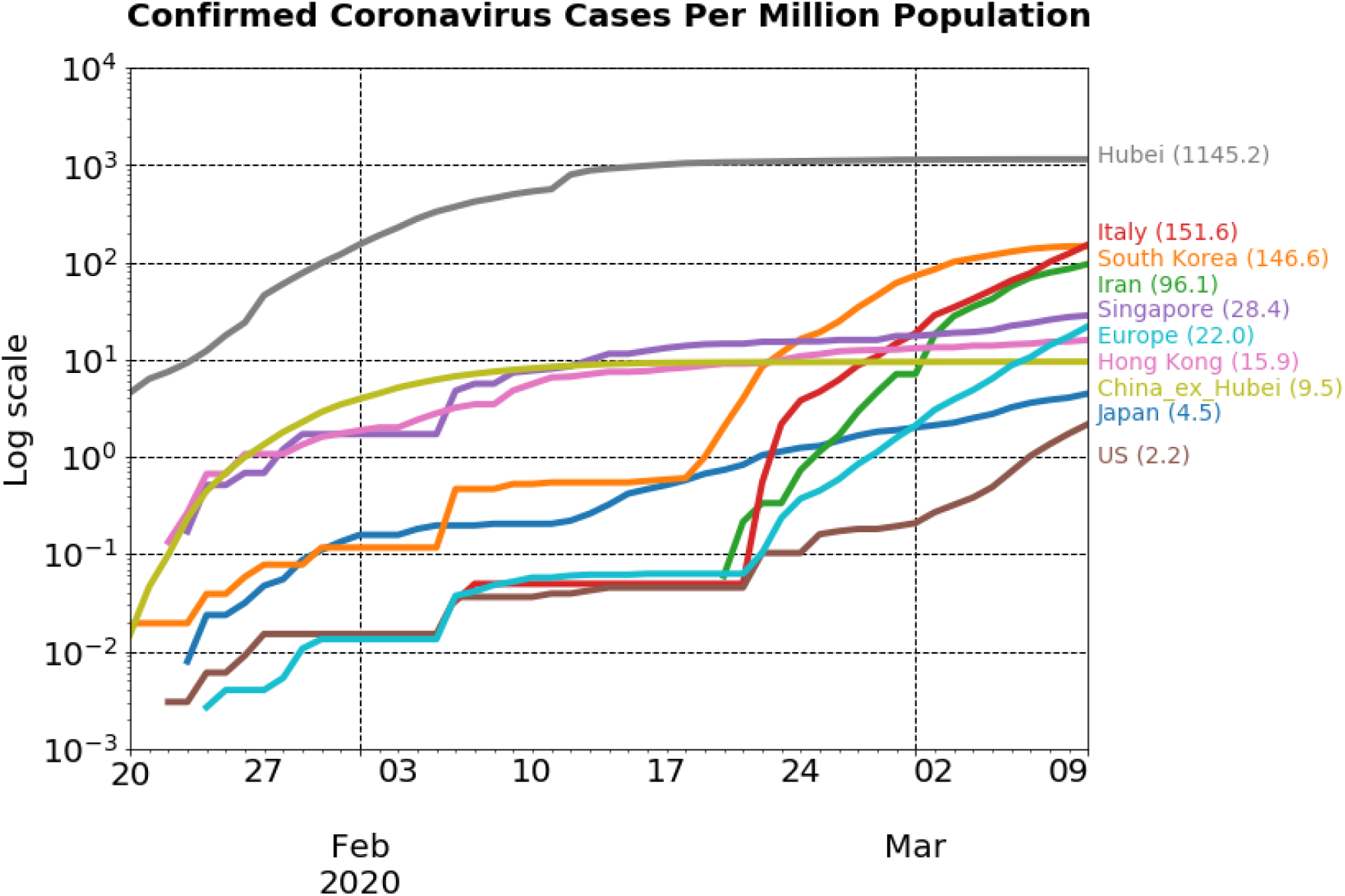
Number of confirmed cases per million people in different countries and regions as of March 10.

Since it is delicate to interpret the absolute number of infected cases, we propose to rather focus on the dynamics, which is more informative as we have presented above. For South Korea, we have shown that it is approaching the ceiling in the total number of infected cases and we do not expect this number to grow significantly in the coming future. If we assume a generalized logistic model for the behavior of Italy, the estimated final fraction of population infected will be 0.15% (95% CI: [0.03%, 0.30%]), which will be much higher than Hubei.

Figure 10 extends the analysis performed for Japan, South Korea, Iran and Italy for the outbreak to the whole Europe, an outbreak apparently mainly initiated by Italy. As of March 10, there are 49 European countries that have reported COVID-19 cases, with Germany, France and Spain each reporting more than 1000 confirmed cases. The statistical analysis shows that the epidemic in Europe is in the early exponential regime and this may continue for a while. At the time of writing, the estimates for the tapering away from a pure exponential growth and the transition to an inflection point and a decay, as obtained from both generalized logistic model and logistic model, are not reliable. There are huge uncertainties simply stemming from the fact that the growth is very close to being pure exponential and does not reveal much additional information.

**Figure 10.**
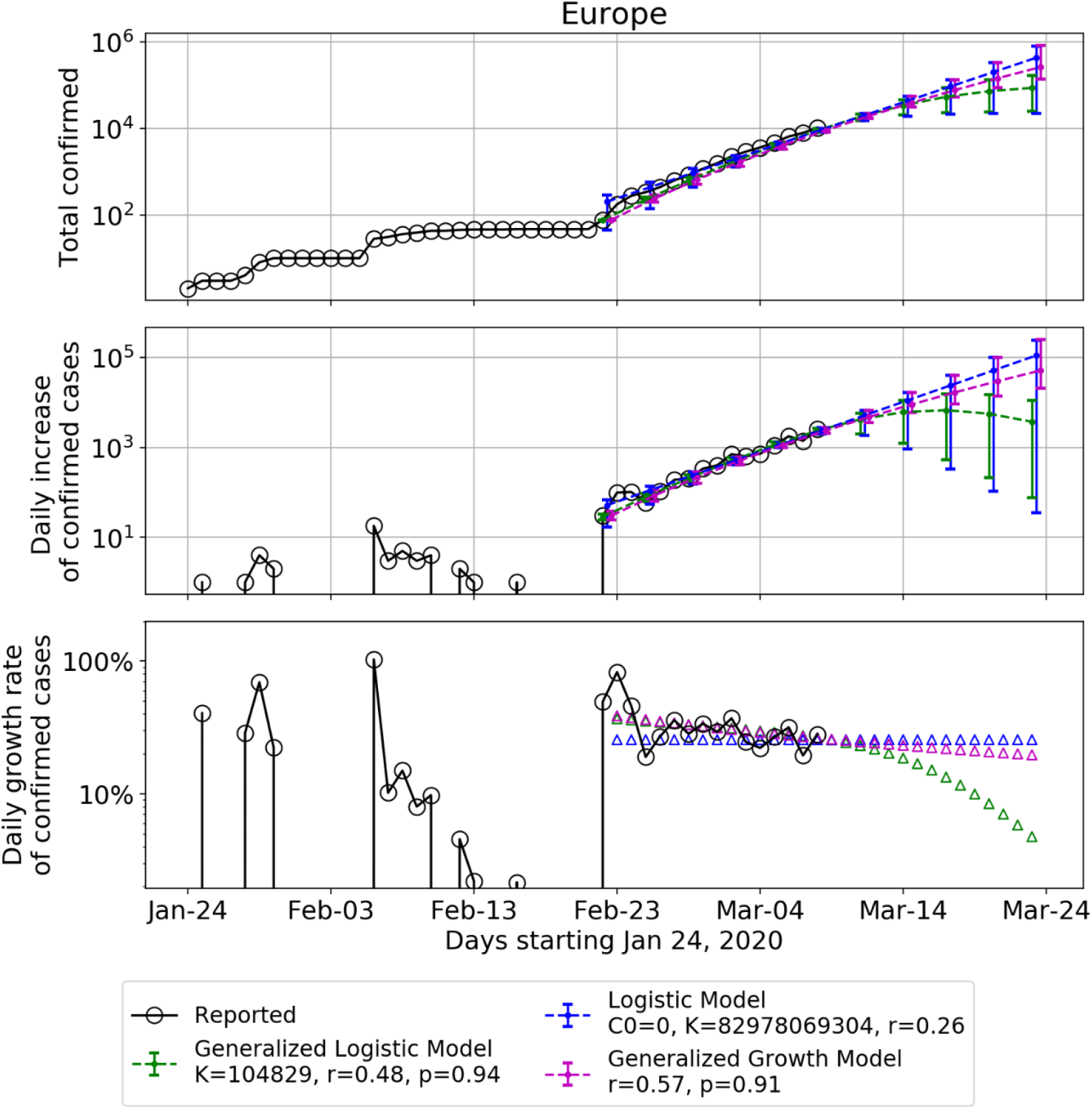
Same as figure 5 for Europe.

Nevertheless, the estimates from the generalized growth model and the generalized logistic model can provide a negative and positive scenario respectively. The generalized growth model assumes that the future growth will continue following the same path as the past weeks, which could be expected as the outbreak is just starting. This scenario will bring the total number of confirmed cases in Europe to 114867 cases (95% CI: [85827, 172995]) in 10 days (Mar-20), and 275451 cases (95% CI: [178462, 545917]) in 15 days (Mar-25). In contrast, the generalized logistic model provides a much more positive (and likely over-optimistic) scenario with the prediction of a total cumulative number of 164219 people infected, corresponding to 0.02% (95% CI: [0.005%, 0.044%]) of the European population of 747.5 million. Given that the generalized logistic model provides the best scenario for Italy with a prediction of a total cumulative number of infected cases of 91719 (95% CI: [16944, 183440]) until the end of the outbreak, it seems that the total number of infected cases for Europe will err towards the upper bound (∼330000) of the 95% confidence interval. Moreover, the reliability of this forecast for Europe can be questioned given the strong heterogeneities between European countries. The situation in the United States is also at the very beginning stage, and may pose significant health and economic risks to the world in absence of serious measures.

## 7. Conclusion

In this paper, we have calibrated the logistic growth model, the generalized logistic growth model, the generalized growth model and the generalized Richards model to the reported number of infected cases in the Covid-19 epidemics from Jan. 19 to March 10 for the whole of China and 29 provinces in China, which allowed us to draw some lessons useful to interpret the results of a similar modeling exercise performed on Japan, South Korea, Iran, Italy and Europe (46 countries aggregated). Our analysis dissects the development of the epidemics in China and the impact of the drastic control measures both at the aggregate level and within each province. We documented four phases: I-early stage outbreak (Jan 19 – Jan 24, 6 days), II-fast growth phase approaching the peak of the incidence curve (Jan 25 – Feb 1, 8 days), III-slow growth phase approaching the end of the outbreak (Feb 2 – Feb 14, 13 days) and IV-the end of the outbreak (Feb 15 – 8 March). We quantified the initial reactions and ramping up of control measures on the dynamics of the epidemics and unearth an inverse relationship between the number of days from peak to the quasi-end and the duration from start to the peak of the epidemic among the 29 analyzed Chinese provinces. We identified the dynamic signatures of the exemplary developments in Zhejiang and Henan provinces and the heterogeneity of the development of the epidemic and responses across various other provinces. We found a strong correlation between the initial and total confirmed numbers of infected cases and travel index quantifying the mobility between provinces.

For countries that are in the middle of the outbreak, we used some useful experience from China to interpret the calibration results from different models, and made future scenario projections. For Japan, the situation is found similar to phase II-III in mainland China and the estimated total number of cases as of March 25 will be1574 (95% CI: [880, 2372]), and 5669 (95% CI: [988, 11340]) by the end of May. However, it took China two weeks to reduce the growth rate from 10% to 1% under extreme containment measures and Japan is still far from achieving this result. Without stronger and fast measures introduced in Japan, we foresee a significant risk concerning the upcoming July 2020 Summer Olympics in Tokyo. For South Korea, we found that it is approaching the ceiling in the total number of infected cases. Both the generalized Logistic model and Logistic model provide similar estimates of the final number of infected cases: 7843 (95% CI: [6594, 9138]) from GLM and 7928 (95% CI: [6341, 9754]) from Logistic model. For Italy, our estimated final fraction of population infected is found equal to 0.15% (95% CI: [0.03%, 0.30%]), which will be higher than Hubei, the epicenter in China of the epidemic. Our statistical analysis showed that the epidemic in Europe is in the early exponential regime and this is likely to continue for a while. This negative but probable scenario will provide 114867 people infected in Europe in 10 days, corresponding to 0.015% European population. The generalized logistic model provides a lower (optimistic) bound, leading to a prediction of a total cumulative number of 164219 people infected, corresponding to 0.02% (95% CI: [0.005%, 0.044%]) of the European population. However, for this forecast to come true, all European countries need to coordinate and fight together to avoid another major outbreak like in Italy. The situation in the United States is also at the very beginning stage, and may pose significant health and economic risks to the world in absence of serious measures.

## Data Availability

The datasets generated and analysed during the current study are available in the Github repository.

https://github.com/kezida/covid-19-logistic-paper

## Declarations

### Ethics approval and consent to participate

Not applicable

### Consent for publication

Not applicable

### Availability of data and materials

The datasets generated and analysed during the current study are available in the Github repository, https://github.com/kezida/covid-19-logistic-paper

### Authors’ contributions

KW and DS designed the research. KW and QW performed the data analysis. All authors wrote, read and approved the final manuscript.

## Acknowledgements

We benefitted from many stimulating discussions and exchanges with Peter Cauwels, Dmitry Chernov and Euan Mearns.

## References

1. Tuite AR, Fisman DN. Reporting, Epidemic Growth, and Reproduction Numbers for the 2019 Novel Coronavirus (2019-nCoV) Epidemic. Ann Intern Med. 2020.(February):2019–20.

2. Zhao S, Cao P, Gao D, Zhuang Z, Chong MKC, Cai Y. Epidemic growth and reproduction number for the novel coronavirus disease (COVID-19) outbreak on the Diamond Princess cruise ship from January 20 to February 19, 2020?: A preliminary data-driven analysis. SSRN. 2020. Preprint at: https://ssrn.com/abstract=3543150

3. You C, Deng Y, Hu W, Sun J, Lin Q, Zhou F, et al. Estimation of the Time-Varying Reproduction Number of COVID-19 Outbreak in China. SSRN. 2020. Preprint at: https://ssrn.com/abstract=3539694

4. Zhang S, Diao M, Yu W, Pei L, Lin Z, Chen D. Estimation of the reproductive number of Novel Coronavirus (COVID-19) and the probable outbreak size on the Diamond Princess cruise ship: A data-driven analysis. Int J Infect Dis. 2020.

5. Li Y, Yin X, Liang M, Liu X, Hao M, Wang Y. A Note on NCP Diagnosis Number Prediction Model. medRxiv. 2020. Preprint at: https://www.medrxiv.org/content/10.1101/2020.02.19.20025262v1

6. Maier BF, Brockmann D. Effective containment explains sub-exponential growth in confirmed cases of recent COVID-19 outbreak in Mainland China. arXiv. 2020. Preprint at: https://arxiv.org/abs/2002.07572

7. Ying S, Li F, Geng X, Li Z, Du X, Chen H, et al. Spread and control of COVID-19 in China and their associations with population movement, public health emergency measures, and medical resources. medRxiv. 2020. Preprint at: https://www.medrxiv.org/content/10.1101/2020.02.24.20027623v1

8. Brandenburg A. Quadratic growth during the 2019 novel coronavirus epidemic. arXiv. 2020. Preprint at: http://arxiv.org/abs/2002.03638

9. Ziff AL, Ziff RM. Fractal kinetics of COVID-19 pandemic. medRxiv. 2020. Preprint at: https://www.medrxiv.org/content/10.1101/2020.02.16.20023820v2

10. Muniz-Rodriguez K, Chowell G, Cheung C-H, Jia D, Lai P-Y, Lee Y, et al. Epidemic doubling time of the COVID-19 epidemic by Chinese province. medRxiv. 2020. Preprint at: https://www.medrxiv.org/content/10.1101/2020.02.05.20020750v4

11. Zhang J, Litvinova M, Wang W, Wang Y, Deng X, Chen X, et al. Evolving epidemiology of novel coronavirus diseases 2019 and possible interruption of local transmission outside Hubei Province in China: a descriptive and modeling study. medRxiv. 2020. Preprint at: https://www.medrxiv.org/content/10.1101/2020.02.21.20026328v1

12. Roosa K, Lee Y, Luo R, Kirpich A, Rothenberg R, Hyman JM, et al. Real-time forecasts of the 2019-nCoV epidemic in China from February 5th to February 24th, 2020. Infect Dis Model. 2020.

13. Roosa K, Lee Y, Luo R, Kirpich A, Rothenberg R, Hyman JM, et al. Short-term Forecasts of the COVID-19 Epidemic in Guangdong and Zhejiang, China: February 13–23, 2020. J Clin Med. 2020.9(2):596.

14. Wu K, Zheng J, Chen J. Utilize State Transition Matrix Model to Predict the Novel Corona Virus Infection Peak and Patient Distribution. SSRN. 2020. Preprint at: https://papers.ssrn.com/sol3/papers.cfm?abstract_id=3539658

15. Lin H, Liu W, Gao H, Nie J, Fan Q. Trends in Transmissibility of 2019 Novel Coronavirus-infected Pneumonia in Wuhan and 29 Provinces in China. SSRN. 2020. Preprint at: https://papers.ssrn.com/sol3/papers.cfm?abstract_id=3544821

16. Chowell G. Fitting dynamic models to epidemic outbreaks with quantified uncertainty: A primer for parameter uncertainty, identifiability, and forecasts. Infect Dis Model. 2017.2(3):379–98.

17. Viboud C, Simonsen L, Chowell G. A generalized-growth model to characterize the early ascending phase of infectious disease outbreaks. Epidemics. 2016.15:27–37.

18. Chowell G, Tariq A, Hyman JM. A novel sub-epidemic modeling framework for short-term forecasting epidemic waves. BMC Med. 2019.17(1):1–18.

19. Chowell G, Hincapie-Palacio D, Ospina J, Pell B, Tariq A, Dahal S, et al. Using Phenomenological Models to Characterize Transmissibility and Forecast Patterns and Final Burden of Zika Epidemics. PLoS Curr. 2016.8.

20. Chowell G, Luo R, Sun K, Roosa K, Tariq A, Viboud C. Real-time forecasting of epidemic trajectories using computational dynamic ensembles. Epidemics. 2020.30(November 2019):100379.

21. Richards FJ. A flexible growth function for empirical use. J Exp Bot. 1959.10(2):290–301.

22. Chan JF, Yuan S, Kok K, To KK, Chu H, Yang J, et al. A familial cluster of pneumonia associated with the 2019 novel coronavirus indicating person-to-person transmission?: a study of a family cluster. Lancet. 2020.395(10223):514–23.

23. Tian Y. 既过年关,也过难关. 人民网 (in Chinese). 2020 [accessed 2020 Jan 25]; https://web.archive.org/web/20200125183422/http://www.xinhuanet.com/politics/2020-01/25/c_1125501347.htm

24. He X. 防控肺炎病毒,”硬核”河南究竟有多硬核?. 每日经济新闻 (in Chinese). 2020 [accessed 2020 Jan 25]; http://www.nbd.com.cn/articles/2020-01-25/1402907.html

25. Lai S, Bogoch II, Watts A, Khan K, Li Z. Preliminary risk analysis of 2019 novel coronavirus spread within and beyond China. 2020.

26. KCDC. 코로나바이러스감염증-19 국내 발생 현황 (3월 8일 0시). KCKC (in Korean). [accessed 2020 Mar 9]; http://ncov.mohw.go.kr/tcmBoardView.do?brdId=&brdGubun=&dataGubun=&ncvContSeq=353431&contSeq=353431&board_id=&gubun=ALL

27. Wikipedia. 2020_coronavirus_outbreak_in_Italy#Spread_to_other_countries_and_territories. [accessed 2020 Mar 9]; https://en.wikipedia.org/wiki/2020_coronavirus_outbreak_in_Italy#Spread_to_other_countries_and_territories

